# The Balancing Act of Academic Clinical Fellows in UK Emergency Medicine: A Qualitative Study

**DOI:** 10.1101/2025.07.17.25331691

**Authors:** Liam Barrett, Thomas AG Shanahan, Rebecca Fish, Virginia FJ Newcombe, Richard Body, Anisa JN Jafar

**Affiliations:** Radcliffe Department of Medicine, University of Oxford, Oxford, United Kingdom; Centre for Clinical Research in Emergency Medicine, Royal Perth Hospital, Perth, WA, Australia; Division of Health Research, Faculty of Health and Medicine, Lancaster University, Lancaster, LA1 4YW, United Kingdom; Department of Medicine, University of Cambridge, Cambridge, CB2 0AW, United Kingdom; Emergency Department, Manchester Royal Infirmary, Manchester University NHS Foundation Trust, Manchester, M13 9WL, United Kingdom; Health Education North West, Manchester, United Kingdom; Division of Cardiovascular Science, The University of Manchester, Manchester, M13 9PL, United Kingdom

**Keywords:** Emergency medicine, Academic Clinical Fellowship, Clinical academia, Trainee experiences, Medical education

## Abstract

**Background:** Emergency medicine (EM) faces workforce challenges in sustaining clinical academic careers. Academic Clinical Fellowships (ACFs) offer protected research time, but little is known about how EM ACFs experience and navigate these posts.

**Methods:** Semi-structured interviews with 20 current and former EM ACFs across 12 UK regions. Interviews were analysed using thematic analysis following Braun and Clarke’s six-phase approach. A mixed inductive and deductive framework was applied. Reflexivity and positionality were addressed through multi-researcher coding and consensus development.

**Results:** Six themes were identified: (1) Elements of surprise — structural ambiguity and unexpected barriers; (2) Unclear direction — limited guidance and inconsistent supervision; (3) Loneliness — professional isolation and detachment from clinical peers; (4) Engagement — enthusiasm linked to research alignment and supervisory support; (5) Repeated generic hurdles — difficulty balancing academic and clinical demands; (6) EM-specific hurdles — reduced exposure to key rotations and limited academic mentorship within EM. Fellows reported uncertainty about training extensions and programme variability.

**Conclusions:** The EM ACF provides valuable entry into clinical academia but inconsistent structures, supervisory support, and clarity in expectations hinder its potential. Standardised induction, tailored supervision, and flexible but transparent pathways are needed. Findings can inform policy, training programmes, and institutional practices aimed at supporting the next generation of clinical academics in EM in the UK.

**Highlights:** *What is already known on this topic:* Academic clinical fellowships (ACF) provide protected research time and are vital for clinical academic careers. However, little is known about how EM ACFs experience these posts.

*What this study adds:* This study reveals that EM Academic Clinical Fellows (ACFs) face significant challenges including unclear structures, inconsistent supervision, professional isolation, and difficulty balancing clinical and academic demands. EM-specific barriers such as limited mentorship and reduced exposure to key rotations.

*How this study might affect research, practice or policy:* To support EM clinical academics, there is a need for standardised induction, clear training pathways, consistent supervision, and tailored mentorship. Addressing these issues can improve retention and development of EM academic clinicians, informing policy and training programme improvements.

## Introduction

The Royal College of Emergency Medicine (RCEM) oversees EM training and research standards in the UK. Its research strategy highlights the need to understand clinical academic posts in EM, identifying the drivers to stay and leave academia, and discovering what encourages engagement in research as key priorities.^1^

Academic clinical fellowships (ACF) in the early stages of specialist clinical training or often the first step into a clinical academic career. The UK’s four nations have distinct academic training pathways. England’s National Institute for Health and Care Research (NIHR) Integrated Academic Training (IAT) pathway (Figure 1) includes 3-year ACF with 75% clinical and 25% research time.^4^

**Figure 1:**
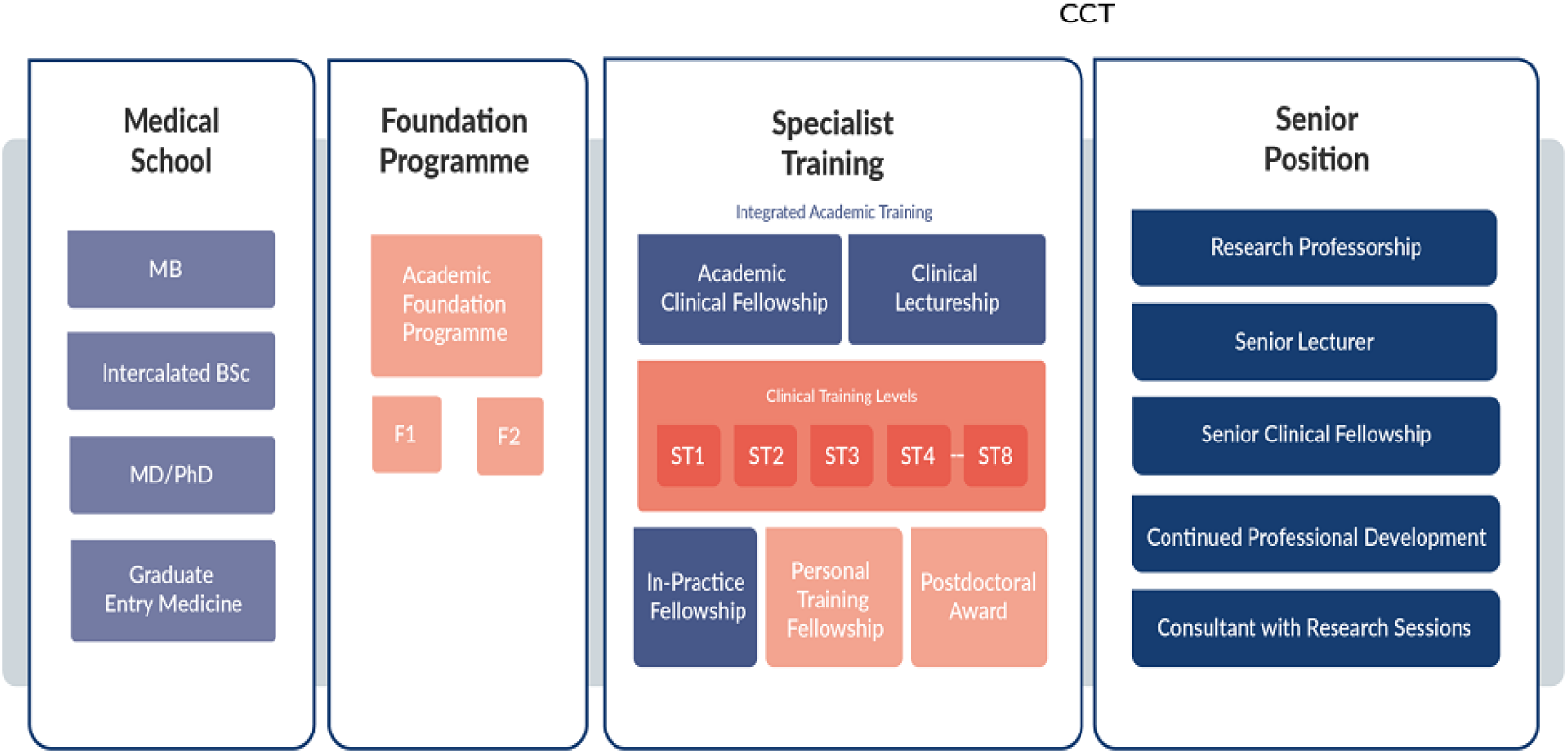
Overview of the Integrated Academic Training pathway in England^4^. Medical school offers the opportunity to do 1-year intercalated bachelor’s or master’s degrees or joint research degrees (PhD/MD) with a medical degree. Academic foundation programme includes the opportunity to do a research block for 4 months, and some offer funding for postgraduate studies. During specialist training there are opportunities to do 3- or 4-year ACFs, a 3-year funded PhD and academic clinical lectureship, which provides up to 4 years of funding during higher specialist training on a 50/50 academic/clinical split. After Certificate of Completion of Training (CCT) there are various competitively recruited senior positions available. Reproduced with permission.

The Wales Clinical Academic Track runs from the beginning of specialist training to Certificate of Completion of Training (CCT). It includes 20% protected academic time and three years out of programme to complete a full time PhD.^5^ In Scotland, the clinical academic track allows for run-through integrated academic and clinical training with a funded PhD and post-doctoral clinical lectureship.^6^ In Northern Ireland, the Queens University Belfast supports specialty trainee year 3 and above to do a two year ACF with 25% protected research time with the aim of applying for PhD funding at the end of the scheme.

A national survey of EM ACFs found 59% of fellows experienced training extensions with wide variation in research time and mentorship and opportunities for funding and postgraduate studies.^8^

This study builds on the national survey and explores in-depth the perspectives of current and former EM ACFs, identifying both barriers and enablers to a successful ACF. We sought to understand how fellows navigate the intersection of clinical and academic roles, how programme design influences experiences, and how improvements could be made to support future cohorts.

## Methods

### Study Design and Setting

This qualitative study used semi-structured interviews informed by a prior national survey^8^. Eligible participants were those that had either completed an ACF (or equivalent in Wales) within the past five years or were currently enrolled in year two or above.

### Recruitment and Sampling

Participants were purposively sampled to ensure representation from all 12 universities that have hosted ACF posts in EM. Recruitment was supported direct invitations, institutional contacts, and social media.

### Data Collection

Semi-structured interviews were conducted by two investigators (LB and TS) via Zoom. Interviews were conducted over a 3-month period in 2023. Audio recordings were transcribed verbatim and anonymised on transcription.

A topic guide, informed by literature and the prior survey, was piloted with a non-EM ACF, and refined following review of the first few interview transcripts by the research team. Interviews explored participants’ experiences of academic supervision, programme structure, challenges, and impacts on career planning.

### Theoretical framework for data analysis

The semi-structured approach was used for the interview with ACFs as it is an effective method for collecting qualitative, open-ended data and allowed interviewers to explore participants thoughts, feelings and beliefs on this topic. ^9^ In addition, the semi-structured approach allowed the research team to explore new concepts not identified in the survey and explain the results behind what was found in the survey. ^9^

All the data (transcripts) from the interviews were thematically analysed by LB and TS and multiple transcripts analysed by AF and RF, using NVIVO software. We used thematic analysis following Braun and Clarke’s six-step process. The six-step process of thematic analysis was followed: 1) data familarisation (all transcripts) as LB and TS interviewed half of the participants each, 2) generating initial codes that were crossed checked by the research team, 3) identification of themes, 4) reviewing of themes, 5) defining and naming themes, and 6) producing the report.^10^ Inductive and deductive approaches were used during the interviews, as an inductive approach allows for the data to determine the themes, but we also used a deductive approach because as researchers we had some preconceived themes based on key areas identified during the survey, which had informed the design of the topic guide. ^11^

### Positionality and reflexivity

The authors have thought carefully about how their opinions, values and conduct shaped how data was generated and interpreted. ^12^ At the time of interview LB and TS were ACFs, with different approaches in their respective institutions. Both LB and TS were in their first year of the ACF at the time and had not completed their programme. Both LB and TS have an interest in widening participation for clinical academics in EM. The positions of LB and TS were disclosed to participants along with their role and experience in the field prior to commencement of the interview.

Given the interviewers were currently participants in the ACF programme it was agreed LB and TS would not interview ACFs from the same institution. The research team consisted of senior clinical academics in EM (RB and VN), both of whom have experience leading NIHR ACF programmes in EM and supervising EM ACFs; an expert in qualitative research methods (RF); and a senior EM resident doctor and experienced qualitative researcher (AJ).

## Ethical Considerations

The project was confirmed by the University of Manchester and the NHS Health Research Authority to be programme evaluation. Informed consent was obtained from all participants.

### Patient and public involvement

Patient and the public were not involved in this study.

## Results

In total, there were 34 eligible participants, 31 of whom responded to a national survey.^8^ We interviewed 20 participants from all 12 locations that have hosted EM ACFs. Interviews lasted between 18 and 60 minutes. The host universities for the EM ACFs represented are listed in Table 1.

**Table 1:** Universities hosting ACFs, included in the study.

| Location of participants |
| --- |
| University of Bristol |
| University of Cambridge |
| University of Oxford |
| University of Exeter |
| University of Leicester |
| University of Hull and York |
| University of Manchester |
| University of Plymouth |
| University of Sheffield |
| Queen Mary University of London |
| University of London |
| Wales Clinical Academic Training (nationally) |

At the time of interview, 55% of the participants declared male gender and 55% reported white ethnicity. The demographics of the potential 34 potential ACFs that were eligible for interview were not captured within the survey. Seven were current ACFs and 13 had completed the programme. Four were current PhD students. Two had completed a PhD. One was an NIHR academic clinical lecturer. One was a consultant.^8^

## Themes

Six themes were identified from the analysis process. These are summarised in Table 2 and expanded in the below text.

**Table 2:**
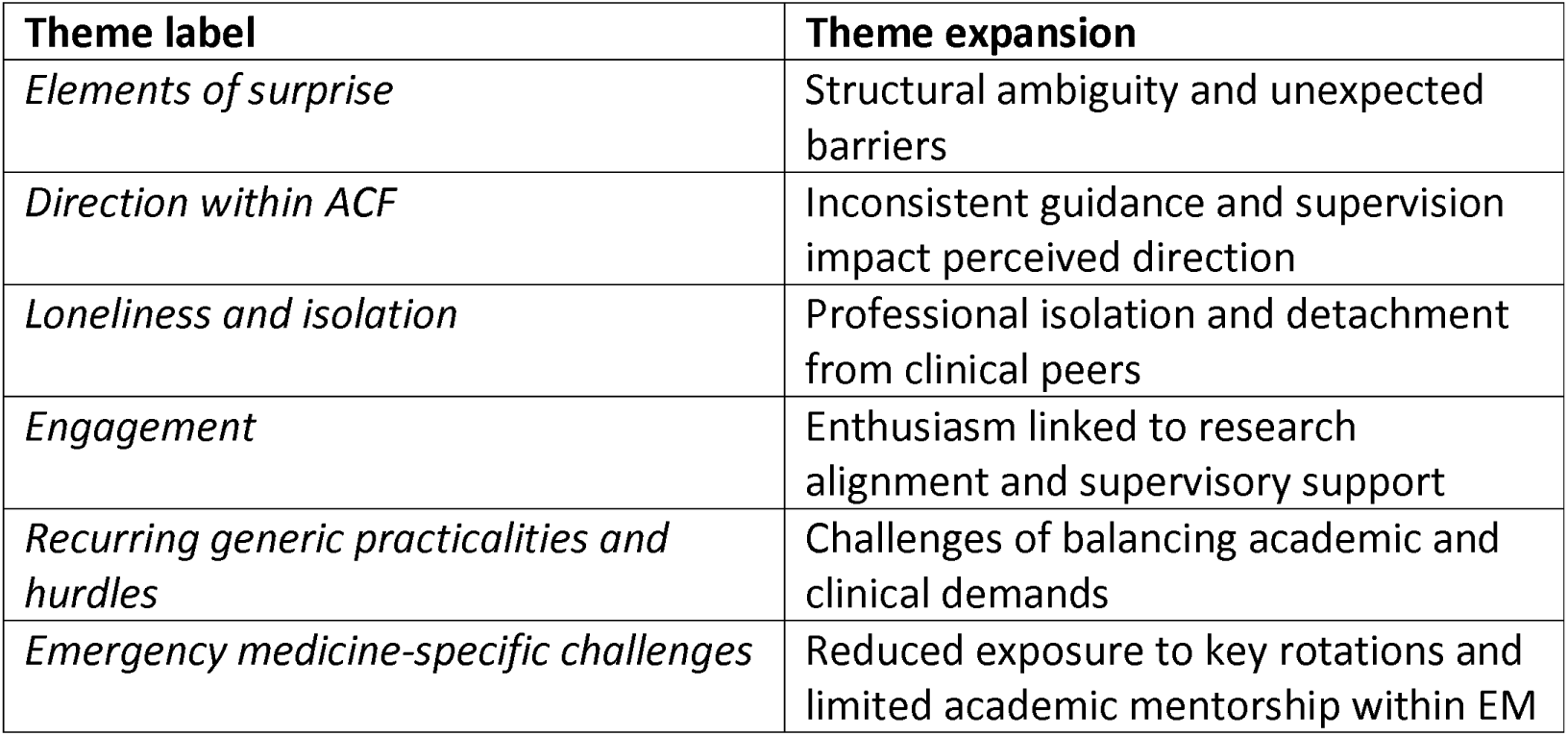
Themes and descriptions identified through analysis of the semi-structured interview transcripts.

### Theme 1: Elements of surprise

The fellowship often seems to surprise its participants with unexpected elements ranging from the flexibility of academic scheduling to administrative complexities and the outcomes required by the ACF program. This significantly impacted both professional trajectories and personal experiences, invoking feelings of uncertainty.

Some specific areas of uncertainty unique to the ACF as compared to non-ACF peer cohort included:

- Administrative hurdles.
- Unclear timing of movement between specialty placements.
- Ambiguous study leave arrangements.
- Unclear expectations for the Annual Review of Competence Progression (ARCP).

Flexibility within the ACF structure, while meant to accommodate individual needs and preferences, often introduced unforeseen complexities and anxiety rather than offering the benefit of flexibility. This uncertainty often led to practical challenges upon returning to clinical duties, due to feeling out of sync.

> *“No one knew I was coming, and no one was aware of me. Even when I rocked up for clinical stuff, there was like one person that knew I was coming. I wasn’t on the rota. I didn’t have access to any IT.”* (interview, 1)
>
> *“A lot of stresses, particularly where rota coordinators and clinical supervisors weren’t necessarily aware of…you being an academic trainee… to be honest it was quite laughable in that my clinical ARCP often didn’t realise that I was an academic trainee.”* (interview, 12)

At times where a trainee had recognised where challenges might lie in the structure of the ACF and training requirements i.e. taking proactive responsibility for where competency hurdles may lie, this was not always accommodated.

> *“The most frustrating was when… I’d asked specifically to do my (ST3) Paediatric block first because…I don’t have any of my competencies signed off… to me that made a lot more sense to try and get that done in six months…then you have got three months of adult, three months of research, and you can top up. But I was put onto adults, so that’s made things a bit more tricky.”* (interview 16)

Being out of sync with clinical peers due to academic commitments commonly affected the fellows’ integration into clinical teams. Missing clinical inductions due to academic schedules often left fellows feeling on the back foot, undermining their effectiveness and confidence in new clinical environments.

Where there was more awareness of the ACF process, there was more visible reported positivity:

> *“My predecessors have done a lot of work to ensure the department understands what is expected of us and what our commitments are.”* (interview, 6)

The unexpected elements of the ACF often disrupted fellows’ sense of stability, revealing the need for clearer expectations and communication, which leads us to explore the critical role of structured guidance within the fellowship.

### Theme 2: Direction within ACF

The direction within the fellowship often leaves participants struggling with ambiguity regarding their roles, expectations, and the structure of the program. This significantly hindered their ability to effectively navigate the fellowship, affecting their independence, sense of achievement, and overall satisfaction.

> *“In the first year, you were just, sort of, told to, well, off you go and be an academic, which is fine if you have a plan, but you could quite easily waste the first three [months] going, ’I don’t really know what to do.’”* (interview, 7)

This absence of structured guidance often placed undue pressure on fellows to self-direct their learning and project management, often without adequate support.

> *“Not really knowing what’s expected right at the start. So, it meant that I took on and did a lot in my time, to the point where I almost ran myself into the ground.”* (interview, 6)

Inadequate planning leads to feelings of being unsupported and unprepared.

> *“Felt very unsupported throughout my entire ACF with no guidance much at all and everything that I tried to do, I just hit a brick wall and…yeah, that’s about it. Yeah, it didn’t spark joy.”* (interview, 17).

In direct comparison, where support was felt to be strong, the response became much more clearly positive.

> *“I just think everyone’s been generally, very supportive and I have enjoyed it, and I would recommend it to future trainees.”* (interview, 20)

The desire for a more structured pathway within the ACF was a recurring account among fellows. Many sought a defined framework to provide a clear route through their fellowship which was simple and clear without being prescriptive: offering enough guidance to foster independence without imposing overly rigid constraints.

> *“I just didn’t have the right combination of time or opportunity to do research in a guided or tutored fashion… If you have got someone who guides you in to doing a research paper or writing a systematic review or shows you the ropes, then I think that makes life a lot easier. I think if you are having to try and learn to do it yourself that is not really a successful way of doing it.”* (interview, 3)

While a lack of structured direction created challenges in autonomy and preparedness, it underscored the importance of clearer guidance, which ultimately links to how feelings of isolation and support were experienced by fellows across different specialties.

### Theme 3: Loneliness and Isolation

Loneliness and isolation represented significant challenges in the fellowship, especially for those in EM. The unique structure of the fellowship and the demanding nature of medical work intensified these feelings.^13^

> *“So, I think being an academic generally is, can be quite lonely, and it depends on what kind of person you are. So, the bit which I found very hard to begin with, was going from doing full time clinical work to suddenly being on your own.”* (interview, 13)

The COVID-19 pandemic significantly underscored the value of community as participants experienced heightened isolation during periods of lockdown and social distancing. This situation exacerbated the already prevalent issue of loneliness, particularly highlighted in contexts such as PhD programs, where isolation is a well-known concern.^14^

Further, the nature of EM involves irregular shift work, which can disrupt personal and social life contributing to a sense of detachment from peers and family who may have more regular hours.^15^

ACF participants seemed to experience an additive effect of the academic plus EM role alongside structural decisions within the fellowship, resulting in increasing feelings of isolation.

> *“I was the odd one out and yet I had no experience with anybody else who was going through what I was going through, I didn’t know who to share it with.”* (interview, 5)
>
> Perceived ineffective supervision also led to isolation and uncertainty.
>
> *“I probably felt quite isolated early on in terms of where to go to for mentorship or career planning.”* (interview, 14)

Being connected to a wider academic community provided some clear mitigation of isolation, and this was repeated in several of the interviews especially given the relatively small community of academics in EM.

> *“Just a way of meeting other…it was mostly other specialties ’because there wasn’t very many emergency medicine academic trainees really. …I met…one of my good friends now… an ACF through (different specialty) and we’re doing a thing together.”* (interview, 11)

The sense of isolation, compounded by the unpredictable nature of academic and clinical demands, emphasizes the need for more robust support networks and connections, making enthusiasm and engagement in the program even more crucial to success.

### Theme 4: Enthusiasm and Engagement

Enthusiasm and engagement in the fellowship were crucial components that significantly influenced the overall experience and success of fellows. Interviewees suggested several elements which are key to this engagement including:

- Passion for projects.
- Effective communication.
- Well-understood setup.
- Tailored supervision.
- Local research interests.
- Tangible training opportunities.

The enthusiasm for specific, well-defined projects significantly enhanced motivation and satisfaction, aligning professional and personal growth with the fellowship’s opportunities.

> *“I think having a supervisor or a series set of supervisors where there was a clear group of projects, I would be able to get involved in that were already established, that I could help get off the ground and be taught how to do research.”* (interview, 3)
>
> *“The most important thing was to be doing research in an area that I felt really passionate about, and also being with a project which had a strong research team that I could plug into, and they were willing to foster my research expertise.”* (interview, 8)

Effective communication within the ACF ensured that fellows felt well-integrated and informed, reducing misunderstandings and fostering a supportive educational environment. One fellow emphasized the importance of proactive communication.

> *“I met with him (supervisor) quite a lot in the run-up to starting my ACF, to plan how we would make it work, and I’ve met with him, you know, quite a few times during ACCS* to make sure that everything was ready for me starting my project.”* (interview, 20)

The quality of supervision significantly impacted the fellows’ engagement and satisfaction. A flexible and supportive supervisor who is genuinely invested can enhance the fellowship experience.

> *“I have a very supportive clinical supervisor who is also an academic so understands the various strains and has given me a lot of latitude especially early on in the programme.”* (Interview, 16)

Exposure to funding for training opportunities and access to networks of like-minded colleagues were also highly valued.

> *“I was very aware that I had opportunities that other trainees don’t have, for example, funding through the master’s and research, funding of time and the ability to negotiate additional study leave off for these training opportunities.”* (interview, 12)
>
> *“I think the network has been really important, like certainly now as a lone researcher I suppose on my PhD, I rely quite heavily on people I met during my ACF and the network within Emergency Medicine, I think is quite important.”* (interview, 19)

There were specific interviews where it was very clear that there were many positive experiences such as open communication and understanding of the ACF in terms of expectations, and local support, allowing challenges to be addressed without any real associated stress.

*ACCS is Acute Care Common Stem which usually refers to the first two years of EM training - the ACF period usually includes this ACCS time.

### Theme 5: Recurring generic practicalities and hurdles

In the articulated journey of ACFs, recurring challenges were a central theme that significantly shaped the fellows’ experiences irrespective of specialty programme.

These hurdles encompassed issues related to supervision, balancing clinical and academic duties, and the inherent structural complexities of ACFs more broadly beyond just EM.

A notable concern expressed by participants centered around the availability and engagement of supervisors, which was considered crucial for guiding ACFs effectively. Despite supervisors often being well-qualified and senior, their limited availability was felt to hinder the fellowship experience.

> *“I think ensuring that there is a programme lead, or several programme leads perhaps would help.”* (interview, 19)

This scenario raises the suggestion that perhaps ACFs could benefit from having more supervisors who might have more time to dedicate to their supervisees: certainly, participants identified Academic Clinical Lecturers (ACLs) who have experienced ACFs as potentially providing that extra support.

> *“I think I’ve been really lucky, ‘because I’ve had two particular people who are ACLs who’ve been unbelievably supportive and really, really, really helpful.” (interview 1) “And I did manage to give that advice to the next ACF who came after me. And it would have been helpful to have that advice before I started, I think.” (interview, 11)*
>
> *“You almost want a clinical lecturer who has been through the process perhaps to take a lead on organising that ACF programme.”* (interview, 19)

The challenge of managing academic activities alongside full-time clinical duties was another significant hurdle. Many fellows found this balance demanding.

> *“I have felt that my clinical competence has suffered. Procedural stuff, decision making.”* (interview, 2)

Additionally, the overarching structure of the ACF often led to extended training durations beyond initial expectations, causing significant personal and professional impacts.

> *“So, I ended up doing another six months of emergency medicine at the end of my EM programme which really annoyed me because it then… my understanding of part of the ACF was that you wouldn’t have to extend your training time.”* (interview, 3).

This extension not only affected fellows’ career trajectories but also contributed to a pervasive sense of stress regarding the future.

The recurring challenges related to supervision, work-life balance, and training time extensions point to the critical need for addressing these practicalities, which are particularly salient within specialties like emergency medicine, where distinct challenges arise.

### Theme 6: Emergency medicine specific challenges

EM trainees encountered several distinct challenges during their fellowship that were considered particularly challenging due to the nature of their specialisation. These challenges notably impacted their ability to balance clinical responsibilities with academic aspirations.

The ACF often coincided with the ACCS period which is heavy in clinical competencies (i.e. when 6-month acute medicine, intensive care and anaesthetics placements are taking place), making it difficult for trainees to meet all requirements. There was a heterogeneity in the different structures of the ACF and the impact it had on future career aspirations. In some places, the academic block was in addition to the usual three years in the ACCS training. In others, the structure resulted in lost time in anaesthetics, acute medicine or intensive care medicine. This was highlighted as potentially impacting on later applications to sub-specialty training, however, it is unclear if this did impact any individual application.

> *“Now I am applying for prehospital emergency medicine (PHEM) training, and one of the criteria is you need six months of ICM, six months of anaesthesia … if I’d have done what the current ACFs are doing… losing clinical time… I wouldn’t have met the essential criteria… and I would have accepted the ACF despite having PHEM career aspirations at the time, I just wouldn’t have known, I wouldn’t have thought about that in advance.”* (interview, 12)
>
> *“I wouldn’t have liked losing clinical time in anaesthetics or intensive care medicine. I think there’s little enough of that as there is. So that would have been a real downside.”* (interview, 2)

The ongoing uncertainty about the completion timeline leads to considerable stress.

> *“I really didn’t know whether the ACF would last just three years and then I would do another year as an ST3 trainee, whether it would last three years providing I met all my competencies and I had to do my clinical training in the same time. That did provoke some anxiety, I suppose, around what do I need to do and when.”* (interview, 14).

The limited availability of senior academic mentors within UK EM significantly affects the fellowship experience depending on the location of the ACF.

> *“I think part of it was also not having the leadership of a chair in emergency medicine… that would have had research projects that you kind of slip into.”* (interview, 3).

Despite these challenges, the ACF itself, is perceived to gain important time and space in and amongst a specialty whose pace doesn’t naturally lend itself to periods of reflection.

> *“When you’re working 48 plus hours average per week, and swapping shifts every other day, you just end up in this zombie-like state that you can’t plan your own meals, let alone think four years ahead of what your project might comprise. So, the biggest point is…the time to reflect on what I want to do and how I want to do it, and then to have access to resources to then develop specific skills or applications of skills.”* (interview 14)

The unique challenges faced by EM trainees within the ACF reflect the broader tension between academic aspirations and clinical duties, highlighting the importance of program flexibility and the need for clear alignment with future career goals.

## Discussion

### Summary of themes and integration with existing research

The transcripts represented a rich experience of a varied cohort of EM ACFs. Whilst some experiences hint at very positive programmes having learned from feedback of prior ACFs, the variability is wide. At a time where there is a growing crisis in the development of clinical academics^10^, a specialty like EM, with a relatively young and fragile research footprint, must embrace the opportunity to reflect on how to keep progressing.

The “surprise” elements in the first theme appeared pervasively through most of the ACF experiences. It is mostly lack of familiarity and understanding amongst the wider clinical teams which engender negative feelings. This is particularly striking where the ACCS programme is being compressed to accommodate academic training, because it represents added pressure to what is already a technically pressured time of gaining competencies, rotating through specific specialties, and making exam progress. This has also been found in UK academic surgical training.^11^ Even without the elements of surprise, this period would have the potential to generate pressure around academic and clinical attainment in a time designed solely for clinical attainment. Therefore, ACFs felt that this should have been explicit a priori.

Furthermore, every attempt to mitigate additional pressure of uncertainty should be made, otherwise we almost set them up to ‘fail’. A one-size-fits-all answer may not be feasible regionally; however, as has been argued by other specialties’ strong leadership and robust transparent pathways would represent a significant improvement in many regions.^11^

The second theme moves into the domain of ‘direction’ within the ACF. Varied experiences of how the progress of the ACF was being measured mean that the direction of the ACF frequently felt uncertain. This has been a highlighted in review of clinical academic training in the UK.^11^ Whilst flexibility and personalization had demonstrable advantages, they do not mutually exclude the idea of having a clear direction. In many accounts, the direction of the ACF experience felt unclear to the point where fellows experienced anxiety as to whether what they were doing was “the right type of thing”, “enough” or even “too much”. Multi-faceted support and supervision may improve this, especially in the context of a framework for each ACF to negotiate agreed goals for milestone reviews. Such a framework may benefit from being specialty-specific, centrally generated and agreed using the experience of those in our interview pool.

The theme of loneliness and isolation is one which features in many aspects of research and academia. There are very predictable junctures in an EM ACFs experience where this will be felt, which creates an opportunity for mitigation. Rotation out of sync with a support system of peers is entirely predictable. Similarly, the limited pool of EM researchers is well known, hence reducing the potential for developing networks. The NIHR incubator for emergency care is developing multi-professional networks for early career researchers and has a national database of mentors, which will help mitigate the difficulty of finding local mentors.^12^

There are clear examples of positive counter-practices to the challenges presented. Excellent and responsive supervision was a clear contributor to the theme on engagement and enthusiasm, as was the alignment of personal research interests with relevant support. The access to training opportunities and exposure to networks were repeatedly cited as firmly positive attributes of an ACF programme.

Relationships, mentorship, networking have been identified in the literature as key interventions to strengthen clinical academic careers.^13^ There was an overwhelming sense that in their recounting of the challenges, our cohort were doing so in the hope of positive change for future ACFs.

The final themes were split into generic practical considerations within ACFs and highly specific EM-related hurdles. The practicalities include formally using designated junior academics in a supervisory role for ACFs because of having more time and a closer relevant experience to being an ACF. Having very few early to mid-career clinical academics in EM does limit the potential for this currently. The balance of clinical and academic work and its impact on clinical training is a perennial challenge which links strongly to the value placed on the future of clinical academics^.11, 13^ If the overall drive is to foster and encourage then the natural shift would be to celebrate fellows’ existence within a specialty programme and hence adapt to help their development.

EM-specific hurdles are pertinent for RCEM to review with respect to how the logistics of ACFs ought to be developed in a way which does not penalise the attainment of either competencies or essential time-based criteria to be able to sub-specialise. The NIHR integrated clinical academic pathway recommends a standard duration of 3 years for an ACF, with 75% of the time devoted to specialist clinical training and 25% to research training.^4^ Meanwhile, the RCEM strategy allows flexibility for the completion of the ACF within 3 or 4 years.^1^ This duration issue is significant for EM ACFs - with proponents and opponents of both lengthened and shorter timescales. Neither ought to be an automatic barrier for those aspiring to be clinical academics but the lack of clear information in the planning of the ACF leads to ongoing confusion about its implications for both the individual’s personal and professional life. Equally the specialty must recognise its pace in comparison to other specialties, such that time to slow down, think and plan is fundamental to the requirements of developing research. All this comes down to whether EM as a professional entity places value on its capacity to protect and continue to produce high-calibre researchers.

### Limitations

We acknowledge the positionality and reflexivity of the research team and their contributions to the study’s design. To minimise potential limitations, we invited a non-clinician qualitative researcher to join the team and ensure the report was contextualised. We also pre-piloted the interview stems with a non-emergency ACF, reviewed transcripts with members of the wider team, and reached a consensus on themes and sub-themes.

Study participants constituted a self-selected population who responded to the invitation to participate in the survey and, subsequently, the interviews. Given the limited pool of fellows, purposive sampling was used to ensure representation from all locations. However, because we did not capture demographic data at the interview stage, we could not determine whether the ACFs interviewed were representative of the overall cohort. Despite this, we managed to interview 20 out of the potential 34 ACFs.

### Implications for policy and practice

The following recommendations are for consideration for EM trainees, supervisors, funders and RECM to improve ACFs in emergency medicine:

1. Standardised ACF induction across all regions to sign post:

◦ Agreed pre-commencement meetings with relevant clinical, academic(s), administrative, HR regional leads.
◦ Formal ACF structure with personalisation where applicable.
◦ Clear outline for the rotational plan.
◦ Clear ARCP expectations.
2. Multiple supervisors (ideally some junior) to mitigate the challenge of any communication breakdown with a single supervisor impacting a full ACF period:

◦ Defined intervals for supervision meetings (with job-planned time for those supervisors).
3. Designated RCEM lead for ACFs to support best-practice-sharing and provide support early to those struggling in their ACFs but for whom local processes seem not to be working:

◦ Local and national mentorship support.
◦ National support group for EM ACFs.

EM ACFs offer valuable protected research time for trainees, but face challenges in structure, supervision and direction. A tailored approach for the individual, alongside improvements in planning and clear information on the structure of the ACF, emerge as key takeaway messages for academic clinical fellows in EM, their supervisors, and funders to consider.

## Supporting information

COREQ checklist

## Data Availability

All data produced in the present study are available upon reasonable request to the authors

## Conflict of interest statement

The authors declare no conflict of interest.

## Authors contributions

LB and TS participated in the research design, conducted the interviews, carried out the data analysis, and led the writing of the manuscript and are joint first authors. RF contributed to study design, data analysis and final write-up. AJ contributed to the research design, provided senior authorship and guidance throughout the project, and participated in writing and revising the manuscript. VN and RB provided project oversight and contributed to the manuscript by suggesting revisions and offering feedback. All authors reviewed and approved the final manuscript.

## Funding

The project was funded by a grant from the Royal College of Emergency Medicine.

## Ethical approval of studies and informed consent

Ethical approval was not required, as the project was confirmed by the University of Manchester and the NHS Health Research Authority to be programme evaluation. Informed consent was obtained from all participants.

## Acknowledgments

All the fellows that gave their time to be involved in the study and share their experiences.

## Notes

### Competing Interest Statement

The authors have declared no competing interest.

